# Metabolomic atlas of dengue virus-infected individuals unveils unique bioactive lipid imprints in the systemic circulation

**DOI:** 10.64898/2026.02.28.26347347

**Authors:** Abdul R. Anshad, Muthuvel Atchaya, Shanmugam Saravanan, Amudhan Murugesan, Siyana Fathima, Ethihas R Mahasamudram, Rajendran Kannan, Marie Larsson, Esaki M. Shankar

**Affiliations:** Infection and Inflammation, Department of Biotechnology, Central University of Tamil Nadu, Neelakudi, Thiruvarur 610 005, India; Department of Microbiology, Saveetha Medical College and Hospital, Saveetha Institute of Medical and Technical Sciences (SIMATS), Saveetha University, Chennai 602 105, India; The Government Theni Medical College and Hospital, Theni, India; Molecular Medicine and Virology, Department of Biomedical and Clinical Sciences, Linköping University, Linköping 58 185, Sweden

**Keywords:** Dengue, metabolomic profiling, high-resolution mass spectrometry, bioactive lipid mediators

## Abstract

**Background:** Dengue virus (DENV) appears to manipulate several cellular metabolic pathways to permit its replication and immune evasion in the host. Here, we employed high-resolution mass spectrometry (HR-MS) to investigate the serum metabolomic landscape of clinical DENV infection.

**Methods:** Serum specimens from primary dengue (n=11), secondary dengue (n=9) samples, and healthy controls (n=10) were used for untargeted and targeted metabolomic quantification on a Waters Xevo G2-XS QTof Mass Spectrometer. The binding potential of selected ligands against DENV NS1, NS3, and NS5 was evaluated. Crystal structures were retrieved from Protein Data Bank and prepared using the Schrodinger’s protein preparation wizard. Based on findings from untargeted metabolomics, we validated certain bioactive lipid metabolites using commercial enzyme immunoassays.

**Results:** Serum metabolomic profiling revealed multiple distinct patterns for primary and secondary dengue versus controls. A consistent peak was observed at 2.06 mins across all samples. Certain bioactive lipid metabolites, such as, lysophospholipids, phosphatidylcholines, phosphatidylserines, and phosphatidylinositols, were detected alongside carnitine fragments, ceramides, diacylglycerols (DAGs), and bile acid conjugates in dengue. Molecular docking showed that DAG consistently exhibited strong binding to all the DENV proteins. Notably, LPC 22:6 showed a selectively strong affinity for NS5. Enzyme validation showed that in the secondary dengue cohort, LPC was significantly elevated than primary and healthy controls (p<0.05).

**Conclusions:** Our investigations of the metabolomic landscaping, unveiled certain characteristic anabolic shift revealing metabolic vulnerabilities in clinical DENV infection, warranting investigations for use as potential biomarkers of inflammation in disease diagnosis and prognosis.

**Author summary:** Dengue is a mosquito-borne tropical viral infection that can range in severity from asymptomatic to life-threatening manifestations. Dengue virus (DENV) hijacks cellular machinery to sustain its survival in the host. Using high-resolution mass spectrometry (HR-MS), we studied the serum metabolomic imprints of dengue infection. The binding ability of selected metabolomic ligands against DENV NS1, NS3, and NS5 was studied. We found several distinct retention patterns for the dengue cases, with a consistent peak at 2.06 min across all samples. Further, several bioactive lipid metabolites were detected in the dengue infected cohort. Our molecular docking studies showed that diacylglycerol, a lipid metabolite exhibited strong binding with all the DENV proteins. We concluded that certain unique lipid metabolomic imprints exist in clinical DENV infection. The identified metabolomic signatures reveal significant potential for metabolomics to elucidate host–virus interactions, contributing to the advancement of antiviral and symptomatic treatments, along with prognostic or diagnostic biomarkers of dengue disease.

## Introduction

Dengue fever, caused by dengue virus (DENV), represents a major threat to public health, especially across the tropical and subtropical regions where the virus appears to remain endemic (Danis-Lozano et al. 2019; Selvavinayagam et al. 2024). Dengue transmission occurs via the bite of female Aedes mosquitoes, particularly *Aedes aegypti* and *A. albopictus* (Mutheneni et al. 2017). Four antigenically distinct DENVs have been identified circulating in the global population i.e., DENV1-4, all capable of causing dengue disease of varying severities (Bashyam, Green, and Rothman 2006; Mustafa et al. 2015). Although these serotypes share ∼65% of their genome, each serotype induces a distinct immune response likely affecting the severity and progression of dengue disease (Bosch et al. 2020).

Dengue disease progression can be influenced by complex interactions between viral, host genetic, and immunological factors (Halstead and Wilder-Smith 2019). Antibody-dependent enhancement (ADE), original antigenic sin, activation of cross-reactive memory T cells, and immune response to dengue virus non-structural protein 1 (NS1) are key drivers of infection, amplifying inflammation and vascular dysregulation, contributing to severe clinical manifestations, particularly during secondary infections with heterologous viruses (Bhatt et al. 2021; Khanam et al. 2022; Palmal et al. 2024). Clinically, dengue diagnosis starts with protean manifestations such as high fever, nausea, and systemic pain (myalgia and arthralgia). Nonetheless, these symptoms and signs are shared across many febrile ailments, and hence diagnosing dengue solely on this attribute is often irrelevant (Yeh et al. 2017). Employing a combination of clinical features rather than individual symptoms and signs increases diagnostic accuracy.

The DENV infection leads to marked changes in the host’s metabolic systems, as the virus manipulates several cellular pathways to enhance its replication and evade immune defences. These affected pathways include autophagy, which assists in the generation of viral replication membranes; β-oxidation, which supplies necessary energy and molecules; and glycolysis, which is often upregulated to meet the biosynthetic needs of the virus. These differential metabolomes can be identified using an untargeted mass spectroscopy to derive the unique metabolomic profile for DENV infection. Others identified a distinct metabolomic profile in primary dengue infection that reflects host responses against the pathogen (Josyula et al. 2024; Voge et al. 2016; Xu et al. 2024). Metabolomics appears to remain a robust method for investigating virus-induced metabolic changes at the molecular level. This approach provides valuable insights into the interaction between host and virus, and shows promise in the exploration of biomarkers for diagnosis and prognosis, as well as identifying new therapeutic targets in dengue infection (Romagnolo and Carvalho 2021). Here, we employed high-resolution mass spectrometry (HR-MS) to investigate the serum metabolomic landscape of clinical primary and secondary DENV infections and validated the findings using commercial enzyme immunoassays.

## Materials and Methods

### Ethics approval

A cross-sectional case-control study was carried out in accordance with the guidelines of the International Conference on Harmonization Guidelines and the Declaration of Helsinki, to investigate the serum metabolomic landscape of clinical DENV infection. The study protocols were reviewed by the Institutional Ethical Committee (IEC) of the Saveetha Medical College and Hospital, Chennai, for necessary approval for conduct of the research (Ref. No. 114/03/2024/Faculty/SRB/SMCH). All the human subjects were adults, and written consents were duly obtained from all the participants.

### Participants and clinical classification

DENV infection was confirmed either through nucleic acid or serological detection method. The recruited participants were classified into primary and secondary dengue using PanBio Dengue IgM (Cat. No.: 01PE20, Abbott, Lake Forest, IL, USA) and PanBio Dengue IgG (Cat. No.: 01PE10, Abbott, Lake Forest, IL, USA) kits where samples were diluted 1:100. A threshold value of 11 and 22 PanBio units were considered positive for IgM and IgG, respectively. Samples with IgM/IgG ratio >1.2 were considered primary dengue whereas IgM/IgG ratio <1.2 were considered as secondary dengue as per published literature (Aggarwal et al. 2024).

### Sample preparation for HR-MS

Serum samples were thawed on ice and treated for extraction of metabolites. For the HR-MS analysis, metabolites were extracted using methanol and chloroform in equal parts based on standard protocol (slightly modified) described elsewhere (McHugh et al. 2018). The chloroform-methanol-serum mixtures were vortexed and maintained at –20°C for 30 min. The samples were further centrifuged at 12000 rpm for 15 min. Later, the supernatant was extracted before use in the downstream HR-MS investigations.

### Untargeted HR-MS analysis

Serum specimens from primary dengue patients (n=11), secondary dengue patients (n=9) samples, and healthy controls (n=10) were investigated using untargeted metabolomic quantification as per standard protocols. HR-MS was performed on a Xevo G2-XS quadrupole time-of-flight (QTof) Mass Spectrometer (Waters, Milford, MA, USA) using ES ion unispray positive mode. Briefly, prior to injection the supernatant was diluted with methanol, and 1 µL mixture was used for injection. The machine oven temperature was set to 120°C, with a capillary voltage of 1 KV. Acquity UPLC BEH C18 column (130Å, 1.7 µm, 2.1 mm X 50 mm) was used wherein the collision energy was set as 10-30 V. The extracts were injected into the system, and a full-scan analysis was performed over a mass range of 100–1600 m/z. The high-resolution data enabled accurate mass measurements, allowing for precise identification of metabolites. Data acquisition integration was done using a MassLynx4.2 software (Waters, Milford, MA, USA).

### Docking studies

Molecular studies were performed to evaluate the binding potential of selected ligands against the DENV non-structural proteins NS1 (PDB ID: 4O6BN), NS3 (PDB ID: 6MO0), and NS5 (PDB ID: 2J7W). All crystal structures were retrieved from protein data bank and prepared using protein preparation wizard in the Schrodinger suite (Schrodinger, LLC, NY, USA). Water molecules above 5Å from heteroatoms were removed, the missing side chains were added, hydrogen atoms were incorporated and the structures were minimized. Ligands were downloaded from PubChem database and prepared using the Ligprep module (Schrodinger, LLC, NY, USA) to generate 3D conformations. The docking grids were generated at the centroid of co-crystalized ligands in NS1, NS3, and NS5. Molecular docking was performed using the glide module in extra precision mode. For each ligand, the best docked poses were selected based on the glide docking score.

### Validation of lipid metabolites by enzyme immunoassays

Lysophosphatidylcholine (LPC) (Cat. No. MBS2700562, myBioSource, San Diego, CA, USA; Sensitivity, 153 pg/mL) and DAG (Cat. No. MBS2700657, myBioSource, San Diego, CA, USA; Sensitivity, 274.142 ng/mL) were measured using commercial ELISA kits as per the manufacturer’s instructions. The level of choline (Cat. No. ab219944, Abcam, Cambridge, UK; Sensitivity, 40 nm) was measured using a commercial Choline Detection Kit as per the manufacturer’s instructions. The samples were acquired on a GloMax® Explorer Multimode Microplate Reader (Promega, Madison, USA). The metabolites concentrations were correlated with various acute-phase proteins, clinical laboratory parameters, grades of dengue disease severity, and platelet counts.

### Statistical analysis

To assess differences in metabolite levels across the severity groups, data was expressed as mean ± standard deviation (SD). The relative standard deviation (RSD %) was calculated as (SD/Mean) *100. RSD was computed within each primary and secondary dengue, and healthy control to access variations within the groups. The overall group variation was also expressed as RSD between the groups. Metabolomic data were processed and analysed using python (v3.10) with pandas, numpy, scikit-learn, statmodels, and seaborn libraries. Data were z score normalized before analysis and multivariate analysis (MANOVA) were performed to access overall metabolomic difference across the cohort, followed by HSD post hoc test to identify pairwise differences with P<0.05 considered significant. PCA was applied to visualize clustering patterns and to determine variance across the principal components.

## Results

### The metabolomic landscaping HR-MS study cohort involved both primary and secondary dengue patients

We recruited 30 participants that included DENV-infected patients (n=20; mean age 31 years) and healthy controls (n=10; mean age 28 years). ELISA investigations revealed 66.6% dengue patients (n=20) were positive for anti-DENV IgM, 33% (n=10) positive for anti-DENV IgG, 10.0% (n=3) positive for DENV NS1. All three NS1 positive samples showed positivity for both DENV NS1 and anti-DENV IgM. Based on the criteria for classification of dengue as per standard guidelines, nine patients were identified as secondary dengue. Viral load estimation was done for 15 samples, of which all were negative for dengue viremia (**Table 1**).

**Table 1.** Cohort characteristics for demography, laboratory parameters and serum analytes.

| Clinico-demographic parameters | Total | Healthy controls | Dengue patients | P value |
| --- | --- | --- | --- | --- |
| Age, in years | 28 (23.2, 36.2) | 27.5 (24, 29.5) | 28.5 (23.8, 40.5) | 0.37 |
| Sex, male, n (%) | 20(66.7) | 10 (100) | 10 (50.0) | -- |
| Dengue category |  |  |  |  |
| DWS- | -- | -- | 11 (55.0%) | -- |
| DWS+ | -- | -- | 8 (40.0%) | -- |
| Severe dengue | -- | -- | 1 (5%) | -- |
| IgM positive, n (%) | -- | -- | 20 (100%) | -- |
| IgM titer | -- | -- | 38.4 (16.4, 72.7) | -- |
| IgG positive, n (%) | -- | -- | 12 (60%) | -- |
| IgG titer | -- | -- | 31.6 (4.9, 73.3) | -- |
| IgM/IgG ratio | -- | -- | 1.6 (0.8, 4.1) | -- |
| 2° infection, n (%) | -- | -- | 9 (45%) | -- |
| NS1 positive, n (%) | -- | -- | 3 (15%) | -- |

**Footnotes:** All data expressed as median (IQR) unless specified. P values calculated by Chi square test for categorical variable and Kruskal–Wallis test for continuous variables. **Abbreviations:** IQR, interquartile range; DWS-, Dengue without warning signs; DWS+, Dengue with warning signs; NS1, non-structural protein 1

### Numerous divergent retention times were documented in the untargeted HR-MS serum metabolomic analysis in dengue patients

Metabolomic profiling of serum samples revealed multiple distinct patterns with retention times (RTs) ranging between 0.45 and 4.05 min. A consistent peak was observed at 2.06 min across all the samples. Of note, certain RT features appeared selectively in specific samples reflecting an inter-sample variability. These findings demonstrate the reproducibility of HR-MS in capturing a consistent metabolic profile, which likely may serve as a novel biomarker requiring further validation.

### Untargeted metabolomic investigation revealed several bioactive lipid metabolites associated with inflammation and membrane permeability

Untargeted HR-MS profiling revealed a plethora of lipid metabolites in serum including several bioactive lipids like LPC, PC and phosphatidylserines (PS). A consistently recurring metabolomics lipid analyte was detected at RT 3.28 mins and m/z 520.4 corresponding to LPC 18:1 (**Table 2**). Furthermore, LPC 18:2 was observed in the same RT 3.28 mins. Lysophosphatidylethanolamine (LPE) 18:2 was also observed among the analytes. PCs such as PC 34:1 and PC 18:1 were detected besides PS and phosphatidylinositol. Other notable metabolites included carnitine fragment, deoxycholic acid conjugates, ceramide, and DAG. Together, several bioactive lipid metabolites such as lysophospholipids, PCs, PSs, and phosphatidylinositols were detected alongside carnitine fragments, ceramides, DAGs, and bile acid conjugates. Principal component analysis (**Supplementary Figure 1**) revealed distinct clustering of metabolomic profiles among the cohorts. The first principal component (PC1) accounted for 48.48% while PC2 explained 18.57%. Healthy controls clustered mainly at the PC1 complex, whereas partial separation was observed between primary and secondary dengue cohorts, with secondary dengue exhibiting greater dispersion indicating metabolomic heterogeneity.

**Table 2.**
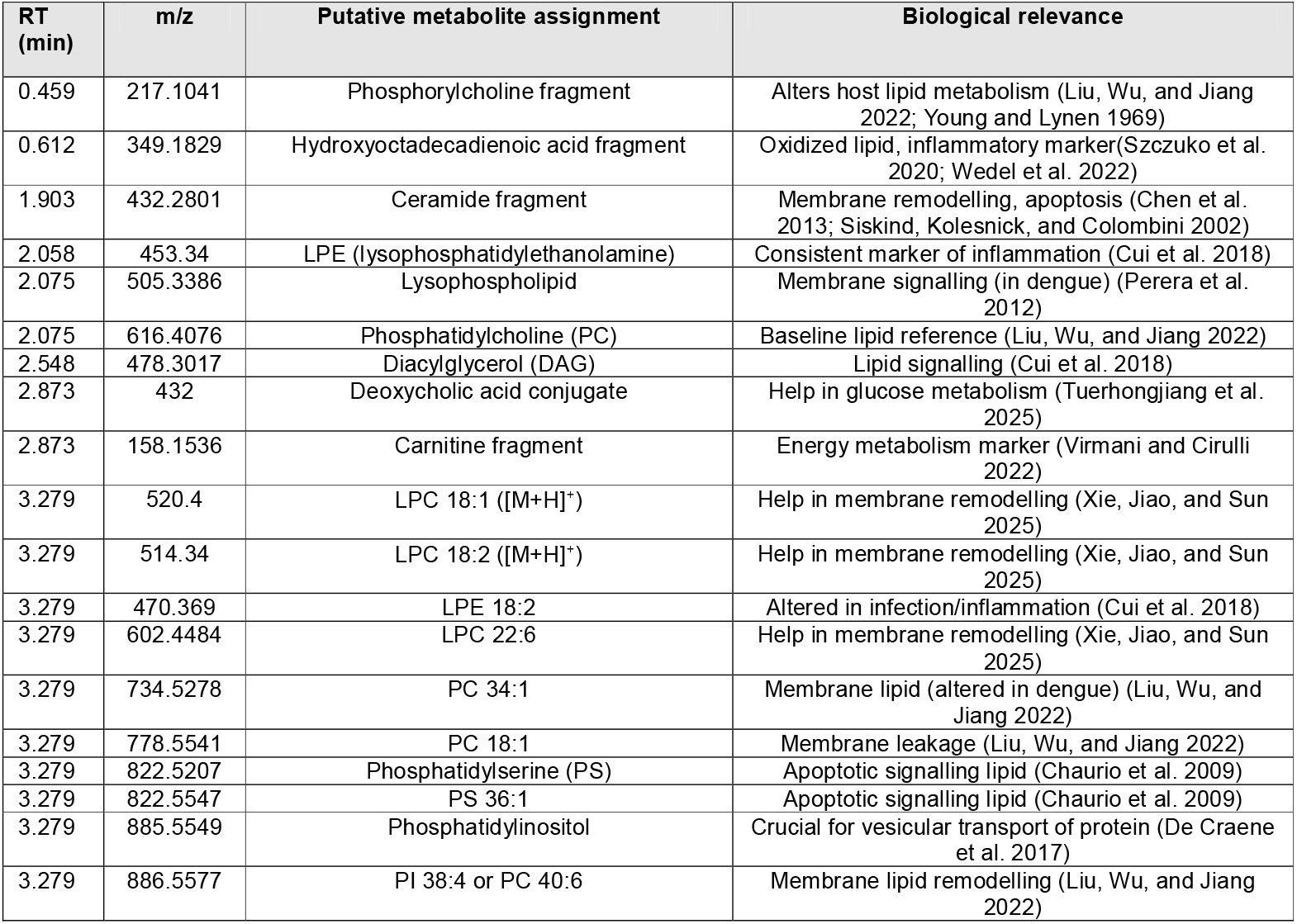
Putative bioactive lipid metabolites detected by untargeted HR-MS in individuals with dengue infection.

### Bile acid-derived deoxycholic acid and several other bioactive metabolites were present across all the study groups

Our HR-MS investigations revealed a distinct alteration in lipid and other metabolite profiles between dengue patients and healthy controls (**Table 3**). We selected a subset of key metabolites (from **Table 1**) for assessing differential expression between the cohorts. PC 18:1 and LPE 18:2 was consistently elevated in dengue groups compared to healthy controls, with a higher variability in secondary dengue cases (RSD: 79.6%). Similarly, LPC 22:6 showed increased abundance in both primary and secondary DENV. Further, certain bile acid derivatives such as deoxycholic acid conjugate demonstrated a uniformly high presence across all the cohorts with minimum variations. In contrast, carnitine fragment exhibited marked fluctuation across the cohorts with high RSD 84.6% suggesting a dysregulated metabolism. Together, the metabolites exhibited distinct patterns of distribution across the clinical groups and healthy controls, collectively suggesting a lipidomic reprogramming characterised by shifts in membrane phospholipids, energy metabolites, and bile acid derivatives in dengue infection.

**Table 3.** Relative distribution of key serum metabolites across healthy controls and DENV patients as measured by HR-MS investigations.

| RT (Min) | m/z | Metabolomes | 1° Dengue (%) | 2° Dengue (%) | Healthy (%) | Dengue (%) | RSD (%) | RSD between 1° and 2° dengue (%) | RSD between cohorts (%) |
| --- | --- | --- | --- | --- | --- | --- | --- | --- | --- |
| 3.279 | 778.5541 | Phosphatidylcholine (PC) | 19.82 ± 12.50 | 17.33 ± 10.36 | 3.00 ± 4.83 | 18.70 ± 11.36 | 67.83 | 9.46 | 53.52 |
| 3.279 | 470.369 | Phosphorylcholine fragment | 50.36 ± 12.20 | 52.56 ± 26.01 | 0.00 | 51.35 ± 19.09 | 86.66 | 3.01 | 66.71 |
| 3.279 | 602.4484 | LPE 18:2 | 49.45 ± 27.81 | 31.56 ± 19.83 | 10.80 ± 15.80 | 41.40 ± 25.62 | 63.21 | 31.25 | 50.12 |
| 2.873 | 432.2383 | LPC 22:6 | 51.45 ± 29.13 | 43.11 ± 27.45 | 14.60 ± 11.36 | 47.70 ± 27.96 | 53.11 | 12.48 | 42.74 |
| 2.873 | 158.1536 | Deoxycholic acid conjugate | 88.18 ± 30.60 | 98.89 ± 3.33 | 48.20 ± 50.86 | 93.00 ± 22.96 | 34.07 | 8.09 | 28.02 |
| 2.548 | 478.3017 | Carnitine fragment | 44.73 ± 31.06 | 35.11 ± 25.64 | 17.10 ± 22.36 | 40.40 ± 28.44 | 43.4 | 17.03 | 35.37 |
| 0.459 | 217.1041 | Diacylglycerol (DAG) | 78.45 ± 23.81 | 82.33 ± 16.09 | 92.40 ± 9.96 | 80.20 ± 20.28 | 8.53 | 3.41 | 7.49 |

### Comparative docking analysis revealed a strong binding affinity of diacylglycerol towards DENV-derived NS1, NS3, and NS5 proteins

Next, we set out to perform molecular docking investigations to assess the binding affinity of three candidate ligands, viz., PC 18:1 (5497103), LPC 22:6 (134747840), and DAG (9547980) against three DENV proteins using the glide module in extra precision mode.

For each ligand, the best docked poses were selected based on the glide docking score. Docking scores were used as predictive indicators of ligand protein affinity, with a more negative value representing stronger interactions. A score □-4 Kcal/mol was considered as the threshold of binding activity, whereas a score lower than -7 Kcal/mol was considered strong binding. Our study showed that DAG consistently exhibited favourable binding across all proteins, highlighting its likelihood to be considered as a target molecule in dengue infection. In contrast, LPC 22:6 showed a selective and strong binding to DENV NS5, but was ineffective against NS1 and NS3 (**Fig. 2A-I and Fig. 3**). Further, PC 18:1 showed a poor performance across all three DENV-derived proteins.

**Figure 1.**
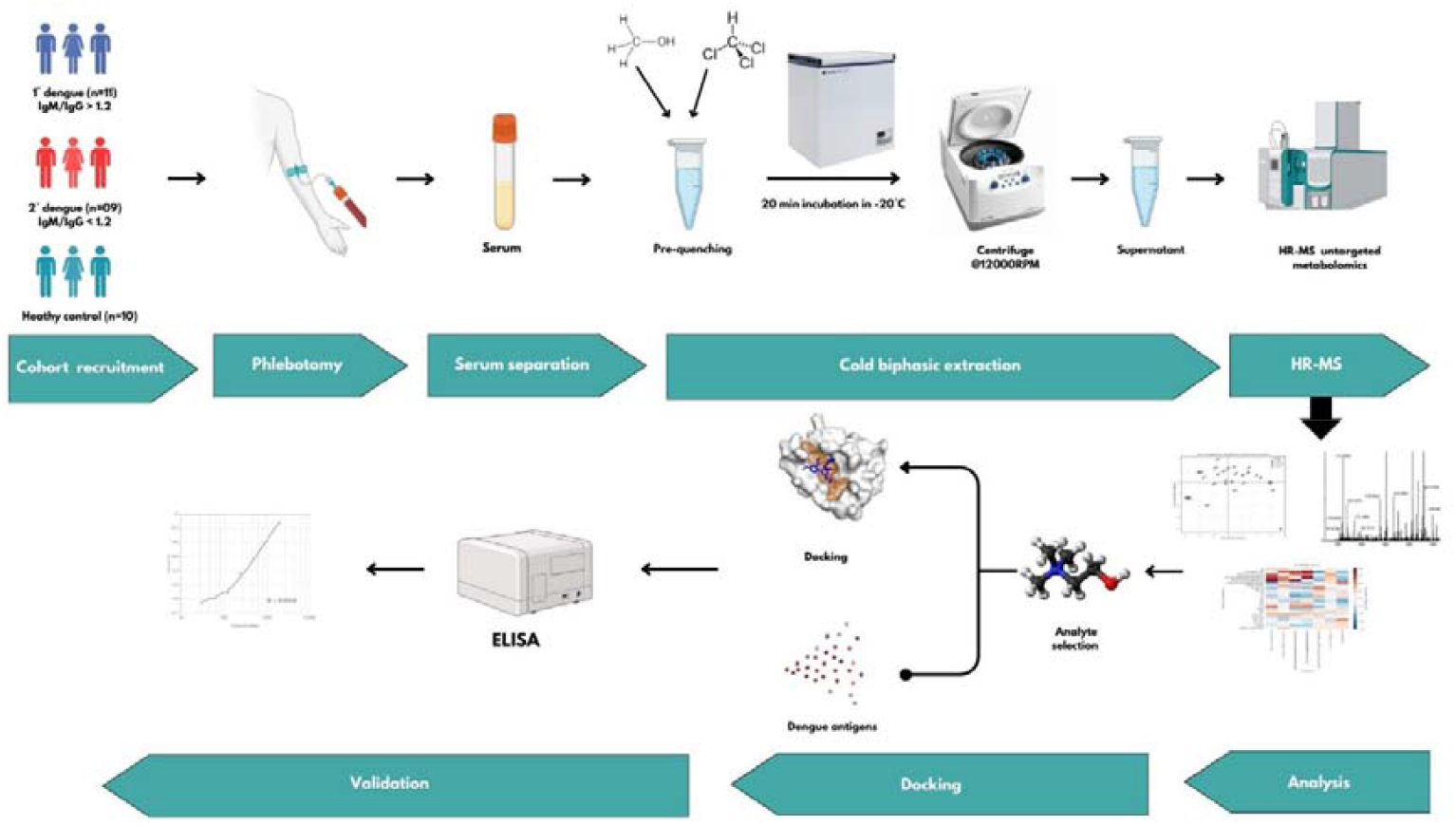
Graphical abstract of metabolomic landscaping in clinical dengue infection. The sera isolated from whole blood drawn from the cohorts (healthy controls, primary, and secondary dengue) were pre-quenched using methanol and chloroform (serum: methanol: chloroform 1:1:1). Untargeted HR-MS was performed on a Xevo G2-XS quadrupole time-of-flight (QTof) Mass Spectrometer (Waters, Milford, MA, USA). Docking studies were performed on a high-performance computing cluster system to measure the interaction between the metabolomes and DENV non-structural (NS) proteins. The serum specimens were collected from patients/volunteers and validated using commercial ELISA.

**Figure 2.**
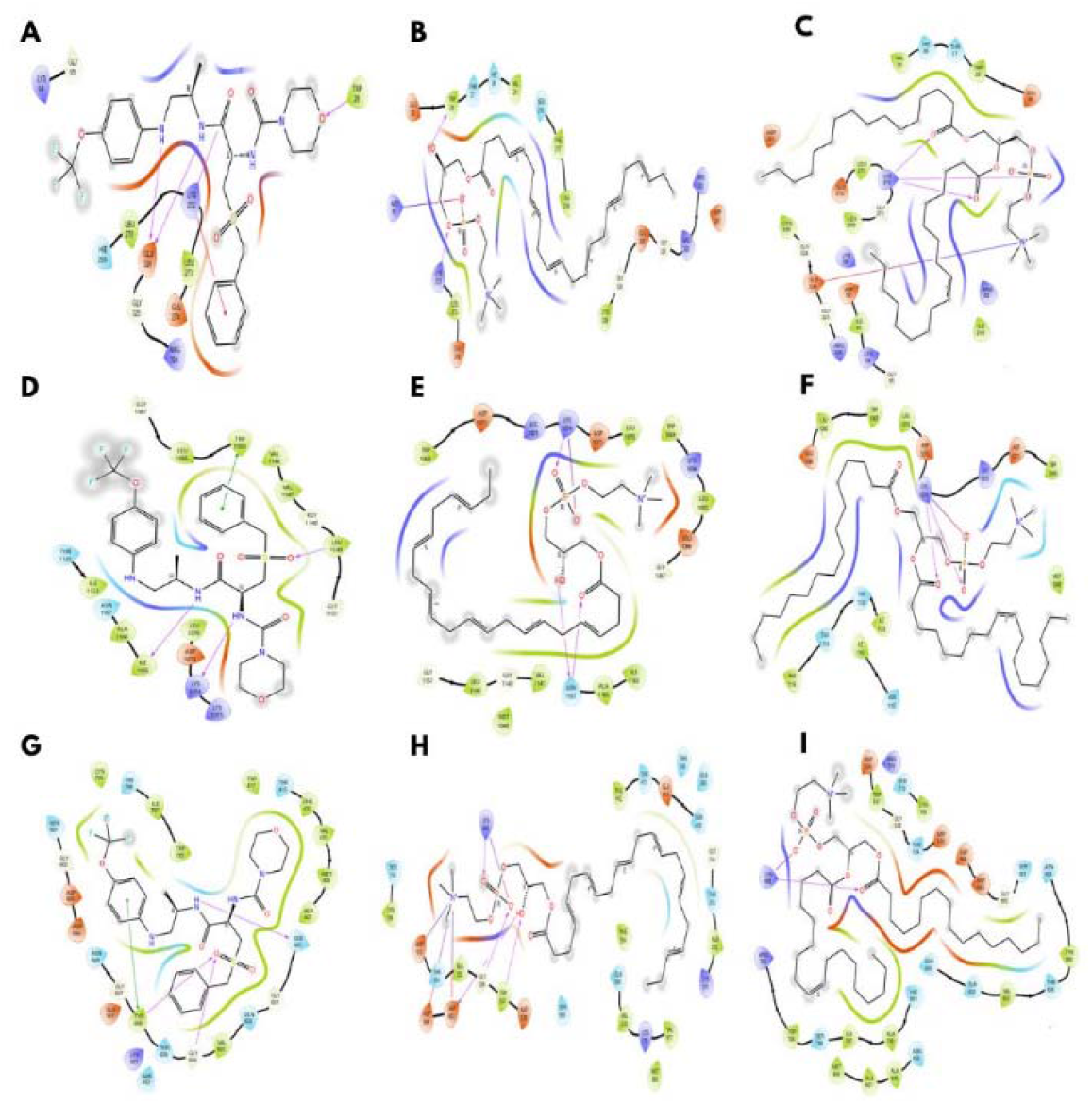
The predicted binding interactions of candidate ligands with DENV-derived non-structural (NS) proteins. Molecular binding structure of selected ligands within the active dengue virus NS proteins. Panel 2A-C depicts the molecular interaction of NS1 protein (PID number: 406B) with a) DAG, b) LPE 22:6, and c) PC 18:1. Panels 2D-F and 2G-I depict the interaction of NS3 (PID no: 6MO0) and NS5 (PID no: 2J7W) with the same metabolite, respectively.

**Figure 3.**
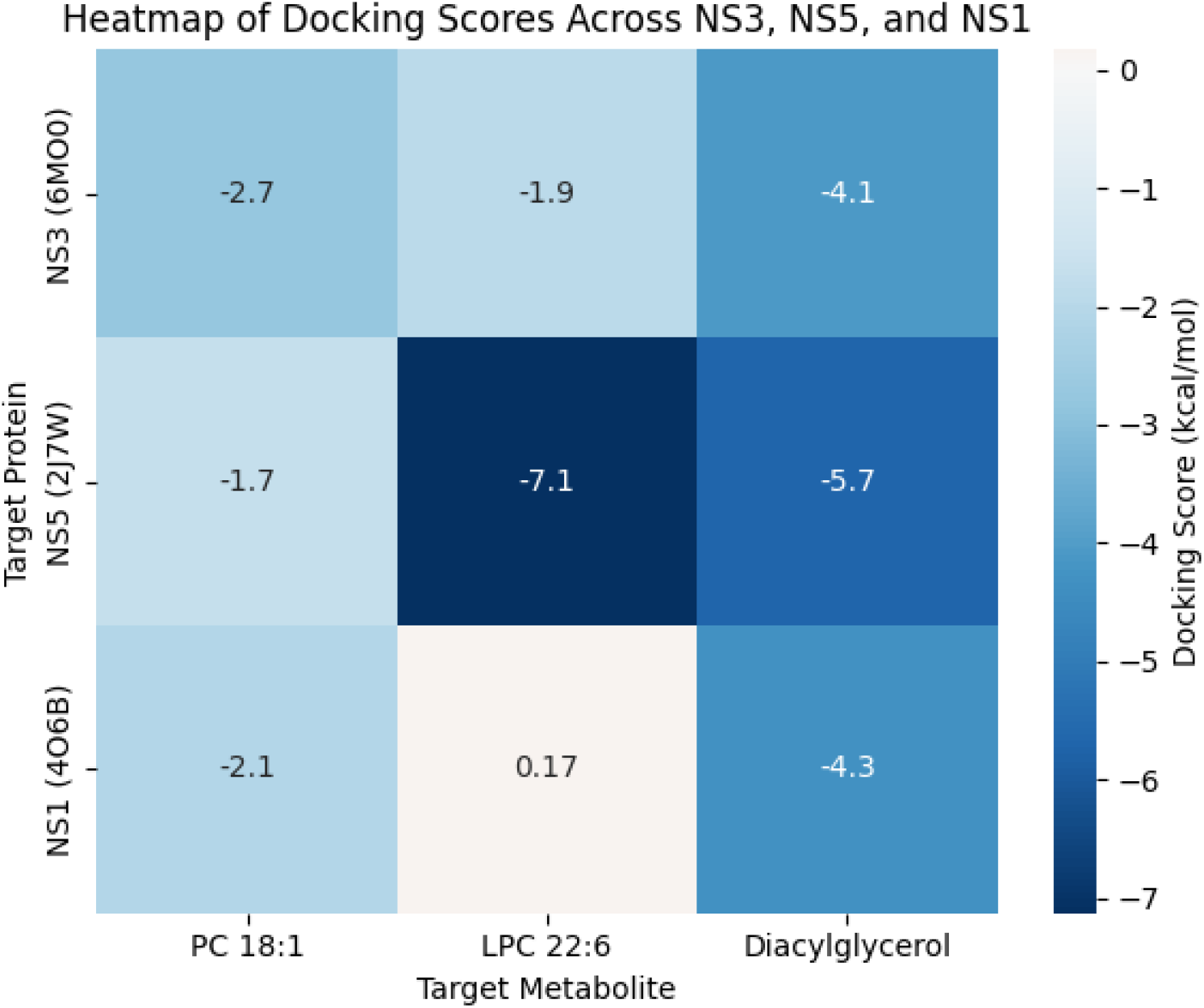
Comparative docking scores of candidate ligands against DENV non-structural proteins. Heat map representation of docking scores (Kcal/mol) for the three candidate ligands against dengue non-structural proteins (NS1 protein, NS3 protease, and NS5 RNA-dependent RNA polymerase). The color scale ranges from dark blue (strong binding, negative scores) to white (unfavourable binding). A lower score indicates higher binding affinity.

### Enzyme immunoassay validation revealed in significant alteration in the levels of lysophosphatidylcholine in secondary dengue infection

Targeted analysis of circulating lipids in serum samples revealed a distinct alteration across the cohorts - primary and secondary dengue, and healthy controls (**Figure 4**).

**Figure 4.**
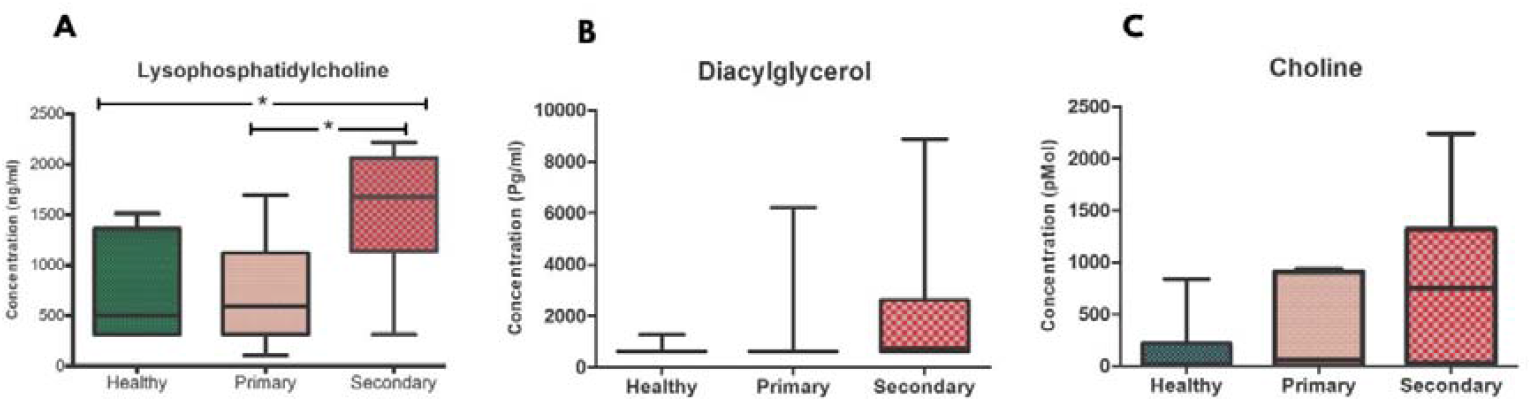
Circulating lipid metabolite levels in healthy controls and dengue patients. Box plot showing circulating **A)** lysophosphatidylcholine (ng/ml), **B)** Diacylglycerol (pg/ml), and **C)** choline (pMol) levels across the study cohorts viz., Healthy controls, primary dengue and secondary dengue. Boxes represent the interquartile range with the median indicated by the horizontal line, whiskers denote the minimum and maximum values. Statistical significance is indicated between primary and secondary dengue. p value <0.05 (significant), where *<0.05, **<0.01, ***<0.001.

Serum DAG levels were elevated in dengue patients relative to healthy controls, with the primary cohort showing a higher median, while the secondary showed higher inter-individual variability. LPC levels were also higher in the dengue cohort, with the secondary dengue cohort showing a significant elevation than the primary dengue cohort (p<0.05) as well than the healthy controls (p<0.05). Circulating choline levels were lowest among healthy controls, with higher variability observed in the secondary dengue cohort.

### Lipid metabolic alterations were associated with clinico-laboratory parameters in dengue infection

Pearson correlation analysis revealed significant associations between selected lipid metabolites and immunological, hematological, as well as biochemical parameters (**Figure 5**). Strong positive inter-correlation was observed among phospholipid species, mainly LPE18:2, LPC 22:6, phosphatidylethanolamine, and phosphatidylcholine molecules (r≈0.7-0.9), indicating a possible phospholipid remodelling.

**Figure 5.**
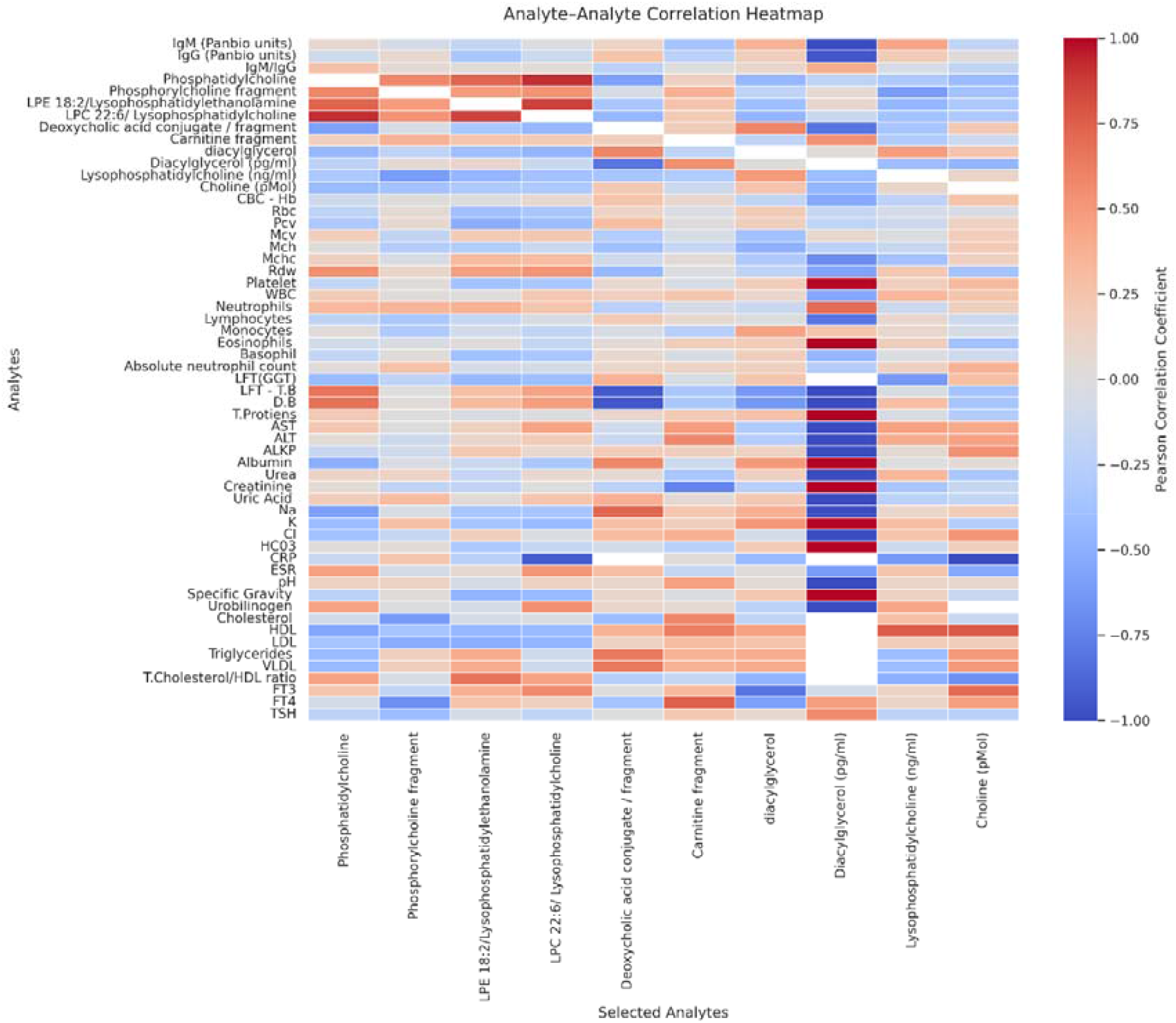
The heat map to depict Pearson correlation coefficients between selected lipid/metabolites and a wide array of immunological, biochemical, and metabolomic parameters. Correlations range from -1 to +1, with red indicating strong positive correlations and blue indicating strong negative correlations. Notable moderate to strong correlations are represented by |r| = ± 0.5.

DAG showed a positive correlation with the liver enzymes and triglycerides. The DAG also showed a negative correlation with the circulating IgG and albumin levels. Deoxycholic acid conjugate had a strong positive correlation with total bilirubin, and a negative association with direct bilirubin. Lysophospholipid showed a moderate positive association with hematological indices, including RDW and leukocytes. Our study also observed a strong inverse association with CRP against several phospholipid metabolites. In addition, thyroid hormones also showed moderate correlation with choline and lysophospholipid.

## Discussion

Dengue diagnosis is classically accomplished via detection of IgM or IgG, or NS1 ELISA, or nucleic acid detection, which often suffers from significant drawbacks (Gyawali and Taylor-Robinson 2017). The most common diagnostic method during the early febrile phase is the detection of NS1, a highly conserved glycoprotein critical for dengue viability (Fisher et al. 2023; Tang and Ooi 2012). Plasma NS1 levels correlate with both viral load and disease severity, and appears during the early stages, but peak only during 3-5 days after symptomatic onset. NS1-based diagnostic kits, with reported sensitivities between 54.2 and 93.4 (Fisher et al. 2023; Tang and Ooi 2012). Besides, anti-DENV IgM are detectable from the third to fifth day of illness, increases over two weeks, and may persist for several months. Nonetheless, although IgM supports diagnosis, definitive confirmation often relies on testing paired samples to establish rising antibody levels. In secondary infections, IgG rises quickly as early as day 4, making the IgM: IgG ratio a valuable diagnostic indicator, whereas in primary infections, IgG is usually not detectable until after 10 days, making it less suitable for early diagnosis.

All these serology-based ELISA techniques show cross-reactivity across flaviviruses due to conserved antigenic epitopes, resulting in false positives, which can make the diagnosis of dengue challenging. Similarly, RNA-based molecular techniques allow detection of dengue viremia, but only at the initial stages, and also often marred with high cost and varying sensitivity across the serotypes (Fisher et al. 2023; Huhtamo et al. 2010; Romagnolo and Carvalho 2021; Tang and Ooi 2012). These factors necessitate the development of improved biomarkers during the early febrile phase and to monitor changes in disease severity, and metabolomics is one such method that shall provide clues to explore novel early biomarkers of dengue infection would be of enormous value for appropriate triaging of patients for management.

Although constrained by modest cohort sizes, published findings have demonstrated that metabolic profiling is capable of producing clinically relevant results, highlighting the differential expression of metabolites based on dengue severity. Others have investigated the spectrum of bioactive metabolites with a few samples and have identified 14 significantly dysregulated metabolites with excellent discriminatory performance (Josyula et al. 2024), justifying the feasibility of our proposed work. Our study has integrated lipid metabolomic profiling and molecular docking to identify the host metabolic alterations associated with DENV infection. The reproducibility of the HR-MS signal across the serum samples, reflected by the consistent retention time peak at 2.058, highlights the reliability of the approach, which will strengthen the possibility and confidence in using the metabolite as a novel method for diagnosis and prognosis of DENV infection, as well as in monitoring other diseases.

The lipidomic environment of the untargeted HR-MS profiling has revealed a broad array of serum metabolites including several classical bioactive lipid components like LPC, PCs and PSs. A consistently recurrent analyte was detected consistently at the same RT that corresponded to LPC 18:1. Moreover, LPC 18:2 was observed in the same RT. The detection of recurrent compounds of LPC in the current study aligns with previous observations highlighting the role of LPC as a modulator of inflammatory cascades and endothelial permeability, which is a hallmark of dengue severity (Voge et al. 2016). Lysophosphatidylethanolamine 18:2 was observed among the analytes. PCs such as PC 34:1 and PC 18:1 were also identified, suggesting membrane remodelling and PC at the higher masses reflecting a possible membrane leakage (Cui et al. 2018). PS and phosphatidylinositol were observed, highlighting host cell signalling and apoptotic pathways. These findings can be attributed to the apoptotic mimicry reported in the flaviviruses infection to facilitate viral entry and subsequent immune evasion (Amara and Mercer 2015).

The detection of serum metabolites including carnitine fragment, deoxycholic acid conjugates, ceramide, and DAG appears to signify the likely implications of DENV with lipid signalling perturbations. Notably, we observed high deoxycholic acid conjugates across all clinical groups with minimum variability, suggesting its role as a baseline metabolite, and not as a DENV infection-specific metabolite. On the contrary, phosphorylcholine is reportedly elevated in the dengue patients, suggesting a strong association with acute inflammation in dengue (Voge et al. 2016). The upregulation of phosphorylcholine in dengue-infected patients points towards alterations in membrane turnover and structural remodelling to create replication-competent organelles, which has been characterised in flavivirus infection. The PC containing metabolites have been implicated in vascular dysfunction and immune dysregulation, two severe hallmarks of dengue infection, supporting the relevance of this finding to the disease pathogenesis (Voge et al. 2016). Moreover, the LPE 18:2 highlights a potential discriminator between the primary and secondary dengue, where the clinically useful markers are very limited. The elevated presence of the LPE might be linked with early immune activation. The downregulation of LPE 18:2 in secondary dengue warrants further exploration, as it might be due to suppression of LPE by pre-existing DENV antibodies, and it may reflect a lipidomic shift highlighting the heterogeneity of host responses in secondary dengue.

In the study, we observed marked alteration of LPC in secondary dengue, suggesting an enhanced phospholipid turnover and membrane remodelling, likely to be amplified in secondary dengue due to heightened immune activation (Soe et al. 2020). LPC is an active biolipid known in the dengue immune pathway, rendering leukocyte recruitment, endothelial permeability, and inflammatory signalling. Its significant elevation in secondary dengue may therefore reflect intensified host immune responses associated with antibody-dependent enhancement (ADE) and inflammatory burden (Carneiro et al. 2013; Knuplez and Marsche 2020). The increase in the DAG observed in dengue patients further supports the involvement of the lipid-mediated signalling pathway, as DAG is a central secondary messenger involved in the protein kinase C pathway and key downstream immune responses (Baldanzi, Ragnoli, and Malerba 2020). The greater variability in DAG levels in secondary dengue might be an indication of heterogeneity in dengue progression, immune activation, as well as hepatic involvement. These findings are consistent with the role of lipid signalling in coordinating antiviral immune surveillance and inflammation (Fonseka et al. 2022).

Our correlation analyses revealed extensive association between lipid metabolites and clinico-laboratory parameters, highlighting the interconnected nature of biolipids and host responses. Strong intercorrelations among the phospholipid metabolites suggest a coordinated regulation of glycerophospholipid metabolism. Furthermore, the association of DAG with liver enzymes and triglycerides is indicative of a link between lipid signalling and hepatic dysfunction, a well-recognised dengue pathogenesis indicator (Hehner et al. 2024; Xie, Jiao, and Sun 2025). This hepatic involvement has been established by the association between bilirubin and deoxycholic acid metabolites (Cui et al. 2018).

The inverse relationship between phospholipids and CRP suggests that systemic inflammation may influence lipid homeostasis, potentially cytokine-driven metabolomic reprogramming (Kurano et al. 2025). Furthermore, the moderate association between lysophospholipids and hematological indices, including RDW and leucocyte counts, are consistent with inflammation-associated changes in erythrocyte membrane and immune activation (Knuplez and Marsche 2020). The observed correlation between thyroid hormones and choline-related metabolites further emphasises the systemic metabolomic perturbations (Post et al. 2025). Moreover, evaluating the phosphatidylcholine, deoxycholine conjugate metabolites, as well as quantifying the circulating choline, allowed assessment of pathway-level concordance. Although these metabolites were measured using different analytical platforms, their biological interrelatedness allows integration of targeted lipids and untargeted data to infer coordinated alterations in lipid metabolism during dengue infection. Overall, these results support the concept that dengue infection, particularly secondary dengue, is characterised by a coordinated disturbance of lipid metabolism and may likely be linked to immune activation, inflammation, and organ dysfunction, which needs to be investigated.

We acknowledge certain limitations in the study. Primarily, the sample size was limited, which may constrain statistical power and the generalizability of the study. Larger, multicentric cohorts may help validate the reproducibility and robustness of the lipid profile identified herein. Secondly, a cross-sectional design to explore the temporal analysis of lipid profile throughout clinical phases of dengue infection is required to further substantiate our results. Similarly, certain confounding factors like nutritional status, and underlying co-morbidities likely influencing the metabolite profile could not be excluded.

## Conclusions

The study demonstrates that serum lipidomic profiling, integrated with molecular docking, offers valuable insights into the metabolic alterations in dengue disease. The consistent detection of LPE, PC, PI, and glycerol molecules highlights the importance of metabolites in DENV infection and their role in inflammatory regulation, membrane modelling, and apoptotic mimicry involved in dengue pathogenesis. The LPC 18:2 identified herein as a potential marker discriminating between primary and secondary dengue, needs further validation to incorporate it into the current diagnostic scenario.

Furthermore, the observed alterations in PC have shed some light on host derived marker and its association with vascular dynamics reinforces its role as a potential biomarker for dengue severity.

## Data Availability

All data produced in the present study are available upon reasonable request to the authors.

## Acknowledgements

The authors gratefully acknowledge technical support extended by the High-Resolution Mass Spectrometer Facility, Vellore Institute of Technology, Vellore, India. Prof. Thangamuthu Mohandas, and Ms. Sheersha Pradhan, Department of Chemistry, Central University of Tamil Nadu, Thiruvarur, India, and Prof. P. Ravanan and Prof. D. Siva Sundara Kumar, Department of Microbiology, Central University of Tamil Nadu, Thiruvarur, India, are gratefully thanked for the instrumentation support extended. The authors also acknowledge Ms. Nachammai Kathiresan and Dr. Langeswaran Kulanthaivel, Department of Biomedical Science, Alagappa University, Karaikudi, India, for offering support with the high-performance docking system, and Dr. Sivadoss Raju, Directorate of Public Health and Preventive Medicine, Chennai, India, for extending instrumentation for their research work. The authors also thank all the national and international members of the Infectious Diseases Society of India (www.idsi.org.in) (IDSI), Chennai for extending insightful discussions pertaining to the research.

## Author contributions

A.R.A. and E.M.S. designed the study and were responsible for conceptualization and data curation. A.R.A., S.F., M.L., Y.K.Y., and E.M.S. conducted the analysis and were responsible for methodology, formal analysis, validation, and visualization. A.R.A., S.F., T.M., M.L., M.A., and E.M.S. wrote the first draft of the manuscript. All authors reviewed and approved the manuscript.

---

**Supplementary Figure 1.**
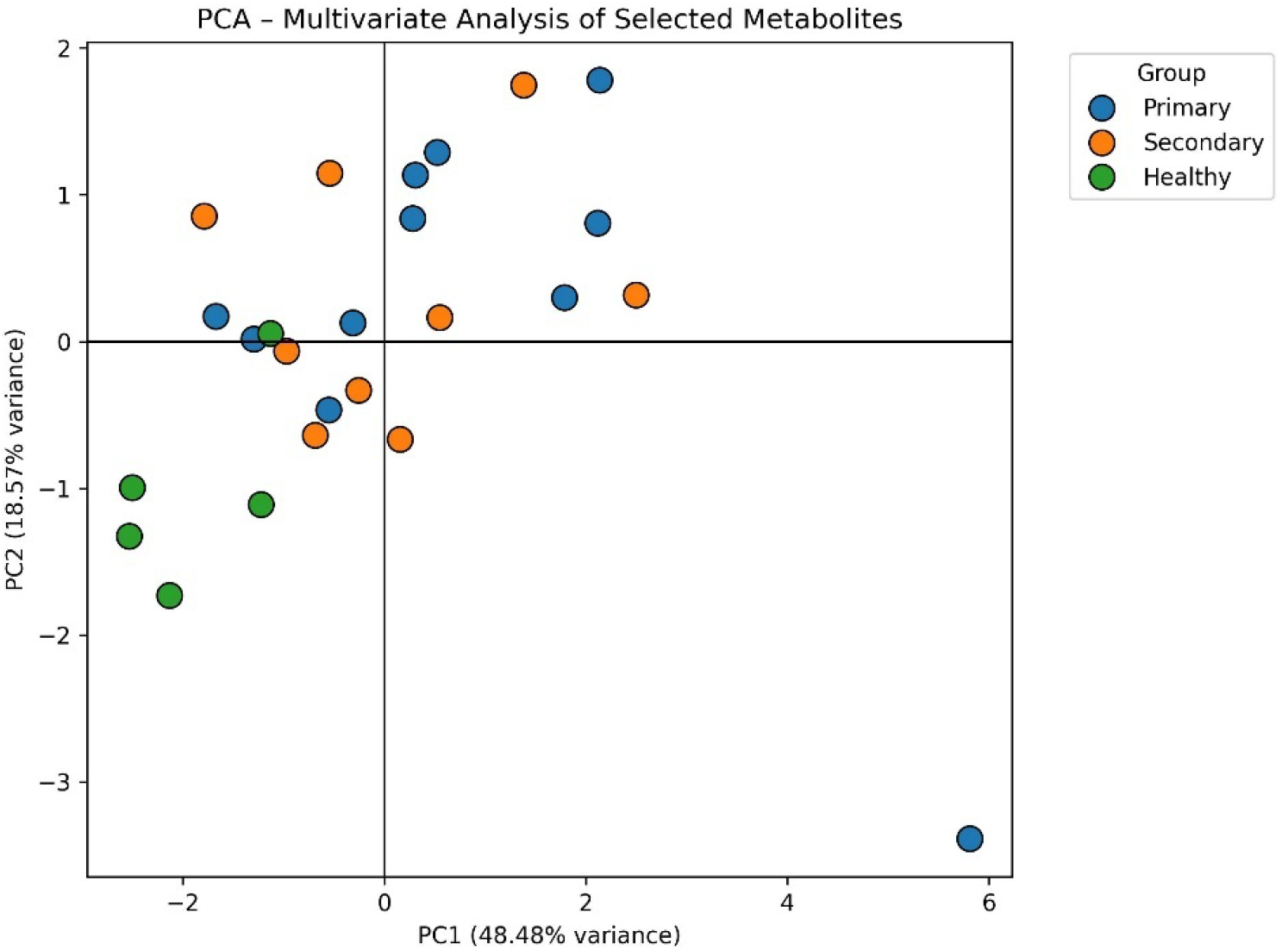
Principal component analysis of metabolomic profiles in dengue. Principal component analysis score plots showing the distribution of metabolomic profiles among healthy (green), primary dengue (blue), and secondary dengue (orange). Each point represents an individual sample. The plot demonstrates distinct clustering of healthy controls and partial separation between primary and secondary dengue.

