## Supplementary Figure 1 for "Metabolomic atlas of dengue virus-infected individuals unveils unique bioactive lipid imprints in the systemic circulation"

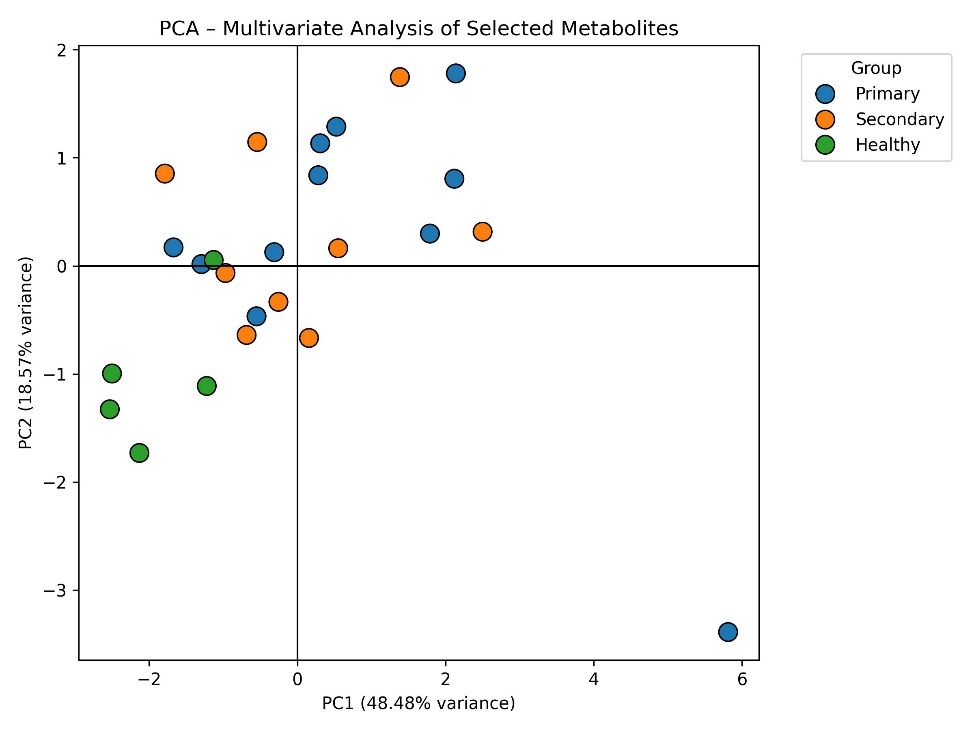

**Supplementary Figure 1. Principal component analysis of metabolomic profiles in dengue.** Principal component analysis score plots showing the distribution of metabolomic profiles among healthy (green), primary dengue (blue), and secondary dengue (orange). Each point represents an individual sample. The plot demonstrates distinct clustering of healthy controls and partial separation between primary and secondary dengue.
